# Vascular growth patterns correlate with histological subtypes in human lung adenocarcinoma

**DOI:** 10.1101/2025.11.28.25340983

**Authors:** Inés Solano-SC, Marta Rodríguez-González, María González-Núñez, Alicia Rodríguez-Barbero, José Manuel Muñoz-Félix, Miguel Pericacho

## Abstract

Lung adenocarcinoma, the most common subtype of non-small cell lung cancer (NSCLC), exhibits diverse histopathological and vascular patterns influencing prognosis and therapeutic response. Angiogenesis, the process of new blood vessel formation, has historically been regarded as the predominant mechanism of tumor vascularization in solid cancers. However, vessel co-option, a non-angiogenic process, has emerged as an alternative strategy, involving the hijacking of pre-existing blood vessels by cancer cells rather than the induction of new ones. Furthermore, vessel co-option has been observed in both primary and metastatic cancers. The aim of this study was to determine the vascular patterns of different human adenocarcinoma samples and establish potential correlations between histological subtypes and microvascular growth patterns. Seventy lung adenocarcinoma samples were classified according to the 2021 WHO classification system, and their vascularization patterns were analyzed using CD31 immunohistochemistry and Weigert-Van Gieson staining. A substantial correlation was observed between histological subtypes and vascularization strategies, with solid basal and diffuse tumors exhibiting angiogenesis, and solid alveolar and lepidic tumors associated with vessel co-option. Additionally, papillary and micropapillary patterns exhibited mixed vascularization, while acinar tumors displayed the highest heterogeneity. The combination of staining techniques improved classification accuracy, achieving successful identification in 92.5% of cases. In conclusion, we present a powerful tool that can be used in lung cancer diagnostics to analyze tumor vascularization based on CD31 and elastic fibers staining. The observed correlations highlight the significance of histopathological assessment in determining vascularization mechanisms, which may optimize therapeutic strategies for NSCLC.

**Highlights:**

- Vascular patterns differ among histological subtypes in lung adenocarcinoma.
- CD31 and WVG staining improves vascular pattern classification.
- Solid basal and diffuse tumors show angiogenesis, whereas alveolar solid shows co-option.
- Papillary and micropapillary patterns display mixed angiogenic/co-option features.
- Vascular profiling may guide NSCLC prognosis and anti-angiogenic therapy effectiveness.

## INTRODUCTION

Within the complexity of cancer, which is the second leading cause of death in developed countries, lung cancer represents the leading malignancy in terms of both incidence and mortality, exhibiting a 5-year survival rate of less than 15% ^1^. Non-small cell lung cancer (NSCLC) accounts for 85% of cases, with adenocarcinomas comprising approximately half of these diagnoses and squamous cell carcinomas representing 30%, though both classifications encompass molecular subtypes of significant heterogeneity ^2^. NSCLC is frequently diagnosed at advanced stages, either localized or metastatic, contributing to poor prognosis due to tumor resistance mechanisms and limited therapeutic efficacy despite recent advancements ^2^. Therefore, there is an urgent need to improve the efficacy of lung cancer therapies through more effective patient stratification.

According to the latest WHO classification, lung adenocarcinoma is graded based on the predominant growth pattern ^3^. There are five main patterns. The one associated with the most favorable prognosis is the lepidic pattern, in which tumor cells grow along the alveolar septa ^4,10,12^. The acinar pattern, characterized by glandular structures embedded in a desmoplastic stroma ^7–9^, and the papillary pattern, defined by fibrovascular cores lined by tumor cells, are associated with an intermediate prognosis ^7,8,10^. Finally, the micropapillary pattern, comprising morules of at least three neoplastic cells free within the alveolar space or within the lumen of tumor glands ^7,11^, and the solid pattern, characterized by sheet-like or cord-like growth where no other defined patterns are recognized, are associated with the poorest prognosis ^3^. The latter has been further subclassified based on cellular arrangement into diffuse, basal, and alveolar subtypes ^6^, and has been subclassified in this way especially in contexts related to tumor vascularization ^6,10,28^.

Despite the growing importance of molecular diagnosis, it is essential to emphasize that a correct histological diagnosis is fundamental. In this regard, the current therapeutic situation of lung cancer demonstrates that the establishment of a treatment requires an accurate histological diagnosis, as many treatable molecular alterations are associated with specific histological subtypes ^13^. Histopathological classification also plays a critical role in prognosis. For example, lepidic-predominant tumors are known to have a better prognosis than acinar or papillary-predominant adenocarcinomas, with micropapillary and solid-predominant tumors having the worst prognosis ^3,9,14^. In addition to their prognostic significance, these growth patterns have also been associated with specific molecular alterations. For example, KRAS mutations have been linked to the solid growth pattern, EGFR mutations to the lepidic pattern, and the cribriform growth pattern, a variant of the acinar type to ALK or ROS1 rearrangements ^2^. Therefore, accurate identification of these patterns is essential.

As with most cancers, lung cancer cells require oxygen and nutrients for their development, and blood vessels play an important role in the delivery of these fuels for cancer cells ^15^. Tumor angiogenesis has been considered for decades as the most important mechanism of tumor vascularization in solid cancers ^16^. For this reason, the study of the cellular and molecular mechanisms involved in tumor angiogenesis has allowed scientists to establish numerous molecules that can target the most important factors in this process: inhibitors of vascular endothelial growth factor (VEGF) or its receptor (VEGFR2), inhibitors of tyrosine kinase activity of VEGFR2, integrin inhibitors, angiostatic molecules or metalloproteinases inhibitors^17^. Several anti-angiogenic agents have been used in the clinical practice for NSCLC treatment such as bevacizumab, ramucirumab and nintedanib ^18^. However, anti-angiogenic drugs have shown numerous limitations and resistance mechanisms ^19^. Among the different mechanisms of resistance are those that allow for alternative tumor vascularization, not based on angiogenesis. Of these non-angiogenic vascularization mechanisms, the most common is vessel co-option ^10^. Vessel co-option is not only a mechanism of resistance but also a non-angiogenic mechanism by which neoplastic cells migrate across the abluminal surface, using pre-existing tissue blood vessels to support tumor growth and metastasis ^4,20^.

Angiogenic tumors have been considered more invasive, more aggressive, and more dedifferentiated than non-angiogenic tumors for decades. However, several authors have demonstrated that tumors undergoing vessel co-option show worse prognosis than angiogenic tumors within the same histological subtype^21^. More recently, it has been proposed that angiogenic tumors have a more favorable tumor microenvironment than vessel co-opting tumors^5^.

Previous studies have highlighted the importance of establishing a correlation between adenocarcinoma growth patterns and vascularization patterns and have proposed preliminar correlations between some histopathological types with both angiogenic and co-option vasculature ^6,10^. However, these studies do not elucidate the histopathological and vascular relationship in all actual histopathological growth patterns, as the classification was updated in 2021 ^3^.

The aim of this study was to determine the vascular patterns of different human adenocarcinoma samples and establish correlations between histological subtypes and vascular growth patterns. The results led to the proposal of a diagnostic tool to more accurately determine tumor vascularization using CD31 labeling and Weigert-Van Gieson (WVG) elastic fibers staining. Additionally, differences in the distribution of non-angiogenic and angiogenic tumors among histological subtypes were observed, enabling the establishment of correlations between specific histological and vascular patterns.

## METHODS

### Human samples

Access to human samples was granted in accordance with the methodology described by the Ethics Committee of the University of Salamanca and within the legal framework of informed consent (Law 41/2002), the practice of biomedical research (Law 14/2007) and data processing and privacy in the national (Organic Law 3/2018) and European (2016/679/EU) context. Seventy human samples of invasive lung adenocarcinoma were provided by the Red de Biobancos de Castilla y León (BEOCyL). All samples were obtained through total surgical resection after an initial determination of stage N0 / N1; therefore, all samples were untreated at the time of resection. The average age of the patients was 67 years (range 46-84), with 45% female and 55% male participants. The samples were processed before delivery using a standard methodology involving dehydration through successive inclusions in increasing concentrations of ethanol (EtOH), clarification in xylene, and paraffin embedding. Once the blocks were obtained, horizontal and uniform sections of 3 µm thickness were cut using a microtome to obtain tissue sections. As a final step, the sections were collected, spread on slides, and dried for 24 hours at 37 °C.

### Hematoxylin/eosin staining

The samples were deparaffinized in xylol for 10 minutes, rehydrated in a decreasing gradient of ethanol (99% and 70%) for 3 minutes, and in distilled H_2_O for 1 minute. Subsequently, Harris hematoxylin (Casa Álvarez) was used to stain the nuclei of the cells for approximately 20 minutes. This was followed by a wash in distilled H_2_O and a quick differentiation in hydrochloric alcohol (HCl 1% in EtOH 70%) to remove excess of hematoxylin. Then, the samples were placed in a bath of running water for 10 minutes and washed in phosphate-buffered saline (PBS) for 1 minute. Next, an alcoholic solution of eosin (Casa Álvarez) diluted with 100% glacial acetic acid was used for 30 seconds to stain the cytoplasmic structures. Finally, the samples were dehydrated using a gradient of increasing concentrations of EtOH, rinsed in xylene for one minute, and mounted with DPX mounting medium (Casa Álvarez).

### CD31 immunohistochemistry

CD31 immunohistochemistry was used to detect tumor blood vessels. For this purpose, samples were deparaffinized and rehydrated in decreasing concentrations of ethanol. Subsequently, antigen unmasking was performed by heat and with Tris buffer [Tris base 50 mM, EDTA 1 mM, pH 9.0] for 10 minutes. This was followed by slow cooling to avoid heat shock with running H_2_O, and after that time, the samples were washed with hydrogen peroxide (3%) and with PBS. After that, they were incubated for one hour at room temperature and under wet conditions with a blocking solution (3% Bovine Serum Albumin (BSA), 5% Tween®20; Sigma-Aldrich). The samples were then incubated overnight at 4°C under wet conditions with a mouse primary antibody against human CD31 (1:100; Dako, Ref. M0823) in 3% BSA and covered with Parafilm. After the incubation time, 3 washes of 5 minutes each were performed with PBS and the samples were incubated with a horseradish peroxidase (HRP)-conjugated goat anti-mouse antibody (1:200; Bio-Rad, Ref. 1706516) in 3% BSA under wet conditions and at room temperature for one hour. Subsequently, three additional washes were performed with PBS for a total of 15 minutes and incubated with the chromogen Diaminobenzidine (DAB; Abcam) for approximately 10 minutes. Once the reaction occurred, the samples were washed with running H_2_O and incubated for two minutes with Mayers’ hematoxylin (Casa Álvarez). The samples were then washed with running water and PBS; and they were dehydrated using increasing concentrations of ethanol, rinsed in xylene and mounted with DPX (Casa Álvarez).

### Weigert-Van Gieson staining

Weigert-van Gieson (WVG) staining was used to detect elastic fibers and collagen in the lung adenocarcinoma samples. In this instance, the staining was carried out with a kit (Casa Alvarez) composed of reagents A, B, C, D, E, F and G, which were used successively, according to the manufacturer’s instructions. This staining is based on three stains: Weigert’s ferric hematoxylin, which stains nuclei black/brown and erythrocytes in yellow; Van Gieson’s picrofuchsin, which stains collagen pink/reddish; and fuchsin-resorcin, which stains elastin purple/brown. Together, the Weigert-Van Gieson stain provides valuable information about the degree of desmoplasia of the samples.

### Sample analysis

Hematoxylin/eosin–stained tissue sections were obtained from tumor blocks fixed in 10% buffered formalin and embedded in paraffin. Tumors were classified according to the growth patterns defined by the WHO: solid, acinar, papillary, micropapillary and lepidic. Solid subtypes were further subclassified into diffuse, basal, and alveolar.

Samples in which the main tumor component consists of solid sheets and lacks other recognizable patterns of adenocarcinoma were classified as solid subtype. The diffuse pattern is characterized by having no recognizable architectural structure; the tumor cells are scattered throughout the tissue in no order. In the basal pattern, cancer cells are arranged in nests surrounded by connective tissue. In the alveolar pattern, cancer cells grow in the alveolar spaces and may completely fill the air spaces. The acinar subtype has been identified as an invasive tumor with well-formed glandular structures of variable shape and size, usually with a central lumen and within a fibrotic stroma. In papillary tumors, cancer cells are arranged in multiple layers around true fibrovascular cores of variable size, and papillary structures occasionally show psammoma bodies. Micropapillary-predominant adenocarcinoma is mostly composed of papillary tufts without fibrovascular cores. Micropapillae may fold on the alveolar surface, float within the alveoli, and sometimes infiltrate the stroma. In lepidic growth pattern, cancer cells line the airspace side of the alveolar walls. They achieve this space by actively replacing the normal pneumocytes that would normally line the alveolar walls. However, cells do not expand to completely fill the airspace and instead they remain firmly attached to the underlying alveolar basement membrane. The classification process was conducted by two independent investigators, with oversight provided by an expert lung pathologist from University Health Care Complex of Salamanca.

Pictures were taken at 1.25x magnification using an optical microscope (BX-51; Olympus31) to analyze the whole slide and delimit different patterns in each slide (Supplementary Figure 1) as described by Yoshizawa ^22^. A slide may exhibit two different growth patterns, and if each pattern accounts for more than 10% of the slide, as reported by Kuhn ^7^, the slide was classified as two or more independent samples each one with an specific histological pattern. For the analysis of vascular patterns, each histological pattern inside a tumor block was considered an independent sample. Thus, 1,25x magnification images of hematoxylin/eosin and vasculature stains were compared to determine the specific vascularization pattern of each histopathological pattern (Supplementary Figure 2 and 3).

### Statistical analysis

Statistical analysis and graphical representations were carried out with the GraphPad Prism 10 software (https://www.graphpad.com). After classification, contingency tables were generated to analyze the relationship between the qualitative variables, histopathological type and vascular phenotype. Relative frequencies of the samples used were then calculated with a p < 0.05 indicating statistical significance.

## RESULTS

### Histopathological classification of lung adenocarcinoma

The different histological subtypes observed in a cohort of 70 patients with lung adenocarcinoma were analyzed using the WHO Classification of Thoracic Tumours 2021 ^3^. Thus, adenocarcinoma samples were classified into the five histological subtypes according to haematoxyilin-eosin staining: solid (diffuse, basal or alveolar), acinar, papillary, micropapillary and lepidic (Figure 1A-G).

**Figure 1:**
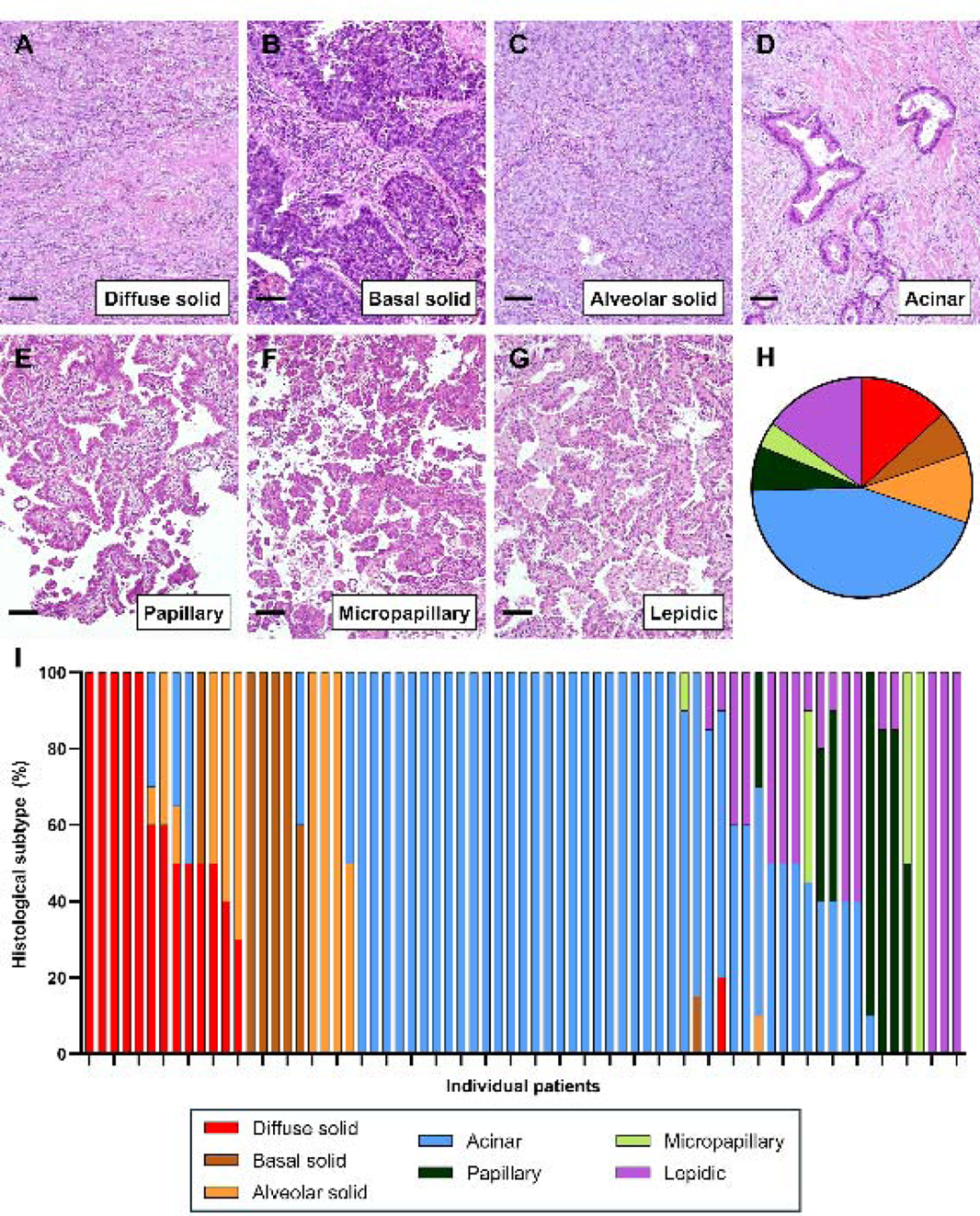
**A-G. Representative images of hematoxylin-eosin staining in lung adenocarcinoma samples.** From left to right: Diffuse solid, basal solid and alveolar solid subtypes; below: acinar, papillary, micropapillary and lepidic subtypes. **H. Histopathologic classification pie chart**. **I. Bar graphs** showing percentages of histopathologic subtypes across individual patients. The patient population was classified and scored (n = 70).

As explained in the Methods section, the entire slide was analyzed, and different patterns were distinguished (Supplementary Figure 1). Therefore, each slide may exhibit one, two or more growth patterns, and if the presence of each pattern exceeded 10% of the sample, the slide was classified as two or more independent samples each one with an specific histological pattern (Supplementary Table 1). Thus, the sample size in our analysis was 99 different histological subtypes from 70 independent tissue blocks. Of the 99 samples, 33% (32 / 99) exhibited solid patterns: 14% (14 / 99) were classified as diffuse solid adenocarcinoma, 7% (7 / 99) as basal solid, and 11% (11 / 99) as alveolar solid adenocarcinoma. Additionally, 47% (47 / 99) displayed acinar pattern, 7% (7 / 99) papillary pattern, 4% (4 / 99) micropapillary pattern, and 16% (16 / 99) lepidic pattern (Figure 1H).

The analysis of the percentage of each histopathological subtype in individual patients allowed for the investigation of which patterns tended to coexist with others and to establish a coexistence relationship. Thus, a bar graph for each individual was plotted to see which patterns were related (Figure 1I). Samples with solid patterns, whether diffuse, basal or alveolar, were observed to coexist with other solid patterns, acinar patterns, and even with lepidic patterns. In contrast, samples with an acinar pattern were associated with papillary, lepidic, or micropapillary patterns. Finally, papillary-predominant samples were frequently associated with micropapillary or lepidic patterns (Figure 1I).

### Blood vessel analysis

To characterize tumor vascularization patterns, CD31 immunohistochemistry was performed to detect blood vessels. The architecture and disposition of tumor blood vessels enabled the identification of an angiogenic growth pattern, which is characterized by an irregular morphology, with a disorganized pattern and an abnormal and chaotic appearance. Additionally, a non-angiogenic growth pattern was identified, where the blood vessels follow an organized alignment according to the alveolar ultrastructure.

Angiogenic growth patterns were observed in the solid, acinar and micropapillary (Figure 2, left panel); and non-angiogenic growth patterns in the solid, acinar, papillary, and lepidic patterns (Figure 2, middle panel). In acinar, papillary, and micropapillary subtypes, non-angiogenic areas coexist with angiogenic areas. These tumors were classified as mixed growth patterns (Figure 2, right panel).

**Figure 2:**
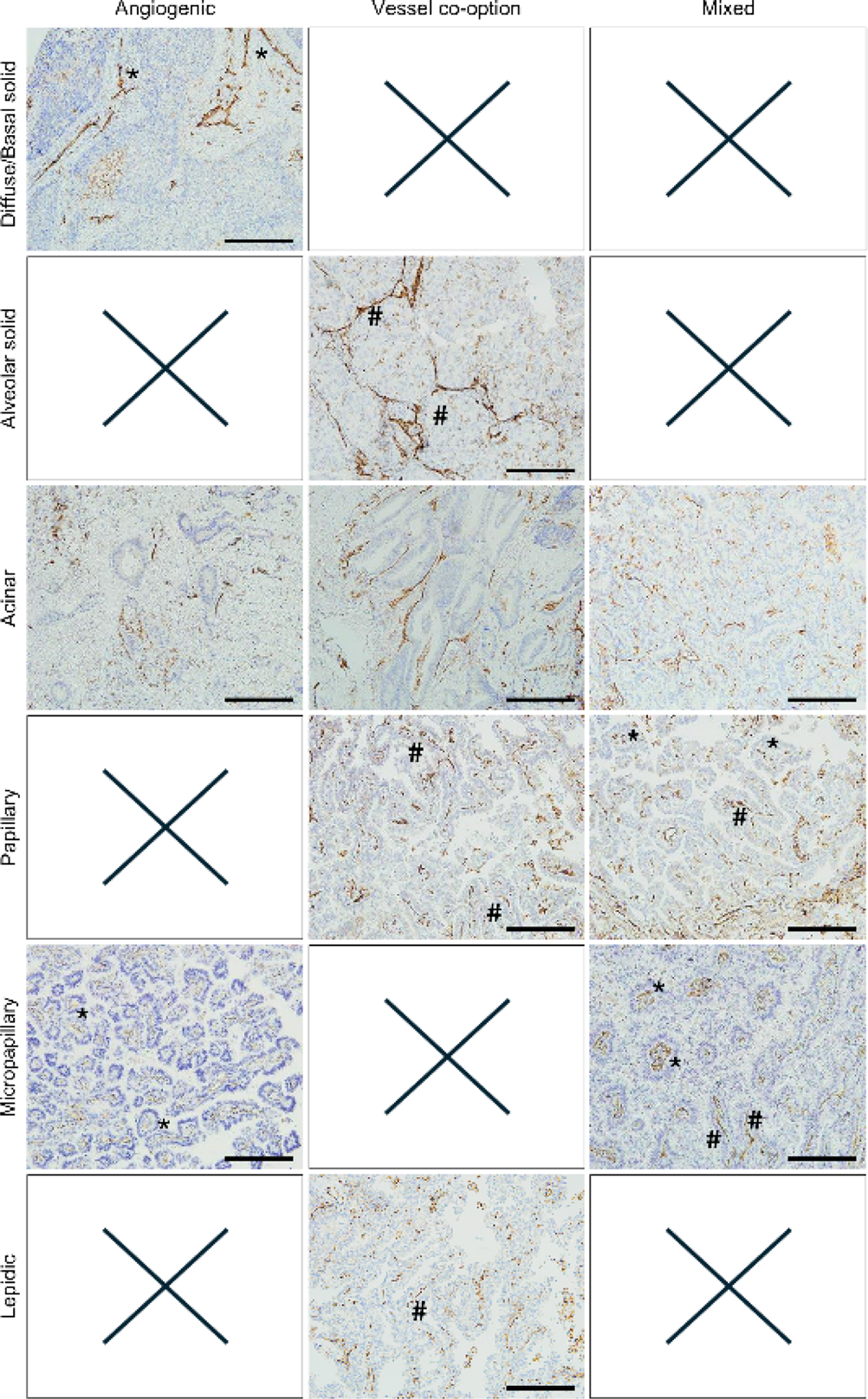
Microvessel patterns (MVPs) identified by CD31 IHC DAB staining (brown). Angiogenic (asterisk), vessel co-option (hash) and mixed patterns (both) are illustrated in the different subtypes: diffuse/basal solid, alveolar solid, acinar, papillary, micropapillary, and lepidic. Panels marked with “X” indicate absence of the corresponding pattern in the subtype and staining. Scale bar: 50 µm.

In diffuse or basal solid pattern, tumor angiogenesis was observed, with tumor vasculature appearing chaotic and disorganized, suggesting that it has arisen *de novo*. Blood vessels in this type of angiogenic tumor also appeared in the desmoplastic space. In this study, as both diffuse and basal solid tumors were angiogenic, they were considered the same subtype. In diffuse patterns, blood vessels were chaotic and intermixed with cancer cells, while in basal solid patterns, most of the vessels were located in the connective tissue area (Figure 2, first row, left panel). In alveolar solid patterns, no angiogenic vessels were observed (Figure 2, second row, left panel). In the acinar pattern, which has irregular glands, new vessels were formed by a angiogenic process, since these vessels lacked an organized structure, and were chaotically organized (Figure 2, third row, left panel). Tumors growing in a papillary pattern did not grow exclusively through angiogenesis (Figure 2, fourth row, left panel). In samples classified as micropapillary, tufts appeared detached within the alveolar spaces while preserving alveolar walls during expansion outside of the main tumor mass. In this case, papillae spreading throughout the airspace predominated, and new blood vessels were formed within these papillae by angiogenesis (Figure 2, fifth row, left panel). No samples described as angiogenic by CD31 staining were found in the lepidic growth pattern (Figure 2, sixth row, left panel).

Conversely, non-angiogenic tumors were characterized by an organized honeycomb structure where cancer cells grew only within the alveolar air spaces, allowing the tumor to co-opt alveolar capillaries contained within these alveolar walls.

In samples classified as solid subtype, non-angiogenic vascularization was also observed, not in the diffuse or basal solid pattern (Figure 2, first row, middle panel), but in the alveolar growth pattern, where the vasculature of the tumor closely resembles the vascular pattern of the normal lung. The morphology of blood vessels was significantly different from that observed in the angiogenic growth pattern. In these samples, alveolar air spaces were filled with cancer cells while alveolar capillaries were preserved. These capillaries appeared thinner likely due to compression by surrounding cancer cells (Figure 2, second row, middle panel). In acinar growth patterns, tumor cells were observed to grow by co-opting pre-existing vessels. These vessels displayed an organized structure and grew around the acini (Figure 2, third row, middle panel). In papillary samples, tumor cells surrounded fibrovascular septa and co-opted alveolar capillaries rather than forming new vessels through an angiogenic process (Figure 2, fourth row, middle panel). Conversely, no tumors with a micropapillary pattern characterized by exclusively co-opted vascular growth were detected. Papillae spreading into airspaces were observed; however, no fibrovascular septa with co-opted vessels were identified (Figure 2, fifth row, middle panel). In lepidic samples, tumor cells proliferated around blood vessels resulting in the co-option of these vessels and the consequent preservation of free alveolar air spaces. In addition, these vessels were noted to be curved and compressed, a characteristic that has been attributed to non-angiogenic vessels (Figure 2, sixth row, middle panel).

Moreover, in acinar, papillary and micropapillary growth patterns, the coexistence of both types of vascularization, angiogenesis and co-option, was identified. More organized vessels surrounding alveoli were observed, as cancer cells can grew along the surface of the alveolar walls to co-opt pre-existing alveolar capillaries. However, new vessel formation through angiogenesis within the alveolar walls with a more disorganized appearance was also noted (Figure 2, right panels).

### Elastic fibers analysis

Some authors have proposed that angiogenic tumors destroy parenchyma, while co-option tumors preserve it ^8,21^. Therefore, Weigert-Van Gieson staining was performed to examine the degree of desmoplasia. Tumor growth patterns were classified according to the preservation of elastin fibers (purple). Angiogenic tumors exhibited either no elastin fibers or disorganized or broken fibers due to destruction of normal parenchyma. Non-angiogenic tumors retained alveolar elastin layers, with elastin appearing as long, dark, tiny fibers surrounding the alveolar stems. In addition, pink collagen filaments maintaining alveolar architecture were observed, demonstrating preservation of the basal lamina of the pre-existing vessels (Figure 3).

**Figure 3:**
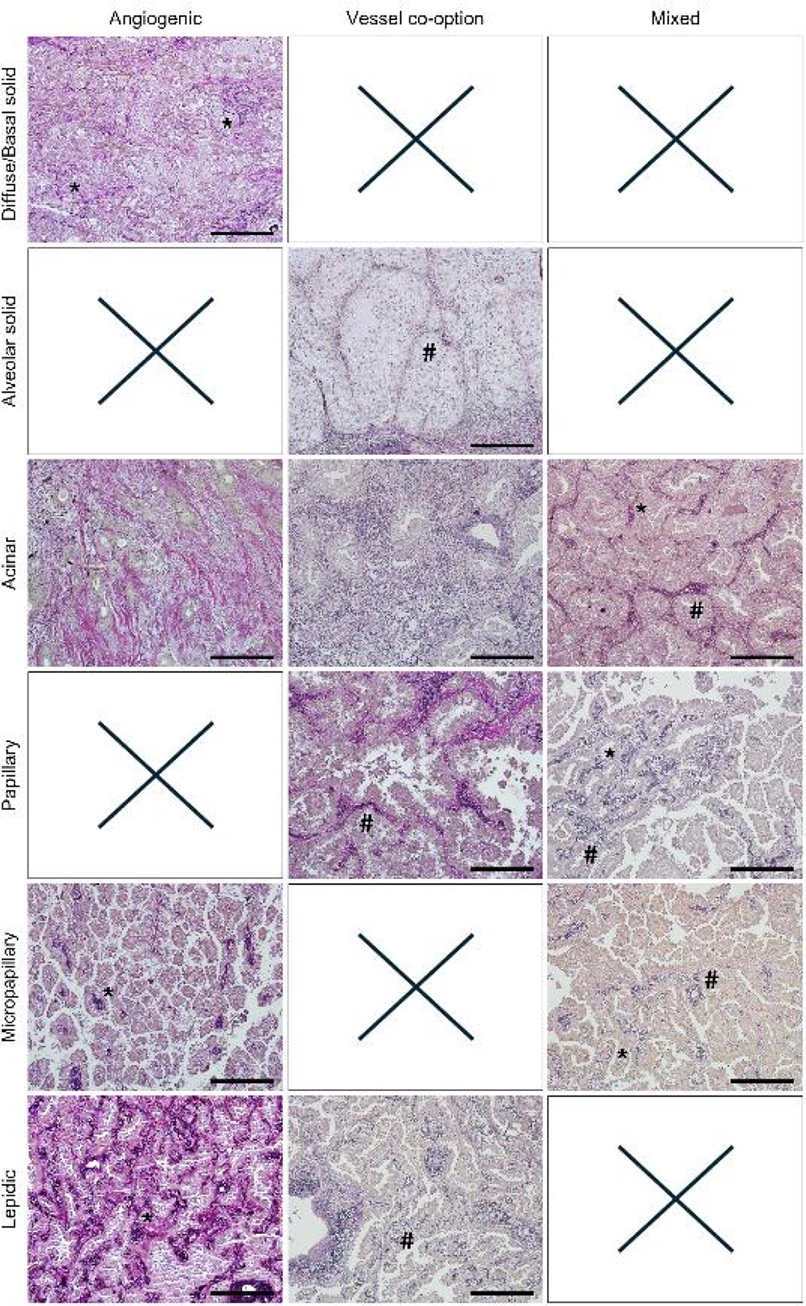
Vascularization patterns observed based on the preservation of elastic fibers identified by Weigert-Van Gieson staining. Angiogenic (asterisk), vascular co-option (hash) and mixed patterns (both) are illustrated across subtypes: diffuse/basal solid, alveolar solid, acinar, papillary, micropapillary and lepidic. Panels marked with “X” indicate absence of the corresponding pattern in the subtype and staining. Scale bar: 50 µm.

Angiogenic growth patterns were observed in the solid, acinar, micropapillary, and lepidic subtypes (Figure 3, left panel); and non-angiogenic growth patterns were identified in the solid, acinar, papillary, and lepidic (Figure 3, middle panel). As in the previous section, in acinar, papillary, and micropapillary non-angiogenic regions coexisted with angiogenic regions. These tumors were classified as mixed growth patterns (Figure 3, right panel).

In diffuse solid patterns, tumor angiogenesis was observed, and the stroma appeared disrupted and desmoplastic, with minimal intact elastic fibers (asterisk) (Figure 3, first row, left panel). Alveolar solid samples did not exhibit exclusive angiogenic growth (Figure 3, second row, left panel). The acinar pattern showed stromal invasion, evidenced by disrupted elastic frameworks with bundles of broken elastic fibers at acinar locations (Figure 3, third row, left panel). Papillary tumors did not grow exclusively via angiogenesis (Figure 3, fourth row, left panel). Some micropapillary pattern samples showed surrounding bundles of black broken elastic fibers in the papillae, indicating of stromal invasion (Figure 3, fifth row, left panel). In addition, some lepidic samples exhibited disrupted elastic framework in areas of stromal invasion (Figure 3, sixth row, left panel).

On the other hand, in diffuse/basal solid growth patterns, parenchymal destruction was observed, but alveolar preservation characteristic of vessel co-option was absent (Figure 3, first row, middle panel). Alveolar solid samples retained continuous black staining of intact elastic fibers in thickened alveolar walls (Figure 3, second row, middle panel). Acinar growth patterns showed unbroken purple elastin fibers surrounding acinar structures (Figure 3, third row, middle panel). Papillary growth patterns were associated with lepidic areas, and stable elastin structures were identified (Figure 3, fourth row, middle panel). On the other hand, no micropapillary tumors with exclusively co-opted growth were detected (Figure 3, fifth row, middle panel). In lepidic samples, tumor cells were observed co-opting blood vessels, with intact elastic fibers resembling normal lung parenchyma (Figure 3, sixth row, middle panel).

In acinar, papillary, and micropapillary patterns, both angiogenesis and co-option areas of preserved parenchyma with stable elastin fibers (hash marks), that identifies co-option, coexisted with desmoplastic regions containing broken or absent elastic fibers, related to angiogenesis (Figure 3, right panels).

### Different histological subtypes are associated with tumor vascular growth patterns

To properly determine the vascular pattern of each sample, the results obtained by CD31 or WVG staining were analyzed together and the data shown in the Supplementary Table 1 were obtained. If a sample was identified as angiogenic by at least one of the stains and classified as either angiogenic or mixed by the other, it was categorized as angiogenic (A). Similarly, if a sample was classified as co-option by at least one of the stains and as mixed or co-option by the other, it was categorized as co-option (C). If both stains indicated that the sample was mixed, it was categorized as such (M). However, if a sample was classified as angiogenic by one staining and as co-option by the other, it remained unclassified (UC). This analysis strategy allowed for the determination of the vascular pattern in 93.4% of the samples, leaving only two samples unclassified. According to the results, 49.05% of the identified samples were angiogenic (A), 34.9% were co-option (C) and 9.43% were classified as mixed (M) (Figure 4A).

**Figure 4:**
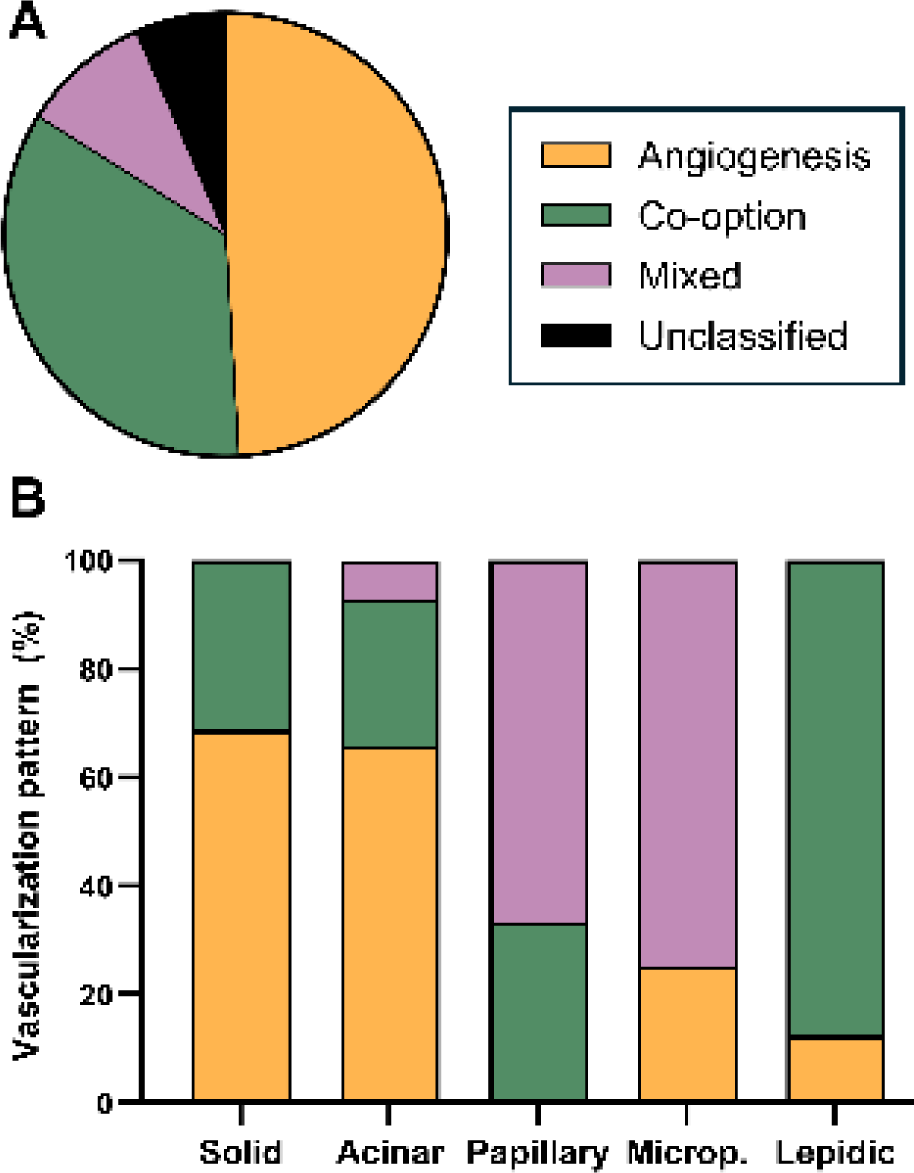
A. Vascular classification pie chart B. Relative frequency of vascularization strategies in the different lung adenocarcinoma growth patterns. The graph illustrates the distribution of angiogenesis, co-option, and mixed vascularization in solid, acinar, papillary, micropapillary, and lepidic adenocarcinomas, highlighting vascularization preferences associated with each histopathological pattern (n = 70).

Most notably, this analysis enabled the calculation of the relative frequencies for each histopathologic growth pattern in relation to vascular patterns, allowing for the determination of their relationship and whether any vascular pattern predominates in any histopathologic pattern. As demonstrated in Figure 4B, solid samples typically exhibited an angiogenic pattern (68.75%), corresponding to diffuse and basal subtypes. However, a proportion of samples (31.25%) demonstrated co-option vascularization, corresponding to alveolar solid growth pattern. Furthermore, it is notable that a significant proportion of acinar samples (65.85%) exhibit angiogenic vascularization, with smaller proportion demonstrating co-option vascularization (26.82%), and only a minimal number exhibiting a mixed vascularization pattern (7.3%). Papillary samples were predominantly classified as mixed (66.66%), with fewer showing co-option vascularization (33.33%). Micropapillary samples were predominantly classified as mixed (75%), with a smaller percentage exhibiting an angiogenic vasculature (25%). Overall, lepidic samples predominantly presented co-option vasculature (87.50%), though two samples exhibit an angiogenic vascular growth pattern (12.50%).

## DISCUSSION

Non-small cell lung carcinoma (NSCLC) is one of the leading causes of cancer mortality worldwide and its treatment continues to face significant challenges due to tumor heterogeneity and the emergence of resistance mechanisms ^1,2^. Consequently, in addition to the ongoing search for new therapies, enhancing the efficacy of existing therapies is necessary, either by avoiding the onset of resistance or stratifying patients to optimize therapeutic decisions.

In the treatment of metastatic NSCLC, one of the first-line therapeutic options is an anti-angiogenic approach, using bevacizumab, ramucirumab or nintedanib in combination with chemotherapy (carboplatin and paclitaxel) and/or immunotherapy (atezolizumab) (National Comprehensive Cancer Network (NCCN; 2024). *Metastatic Non-Small Cell Lung Cancer.* https://www.nccn.org/patients/guidelines/content/PDF/nsclcmetastatic-patient-es.pdf).

Nevertheless, these systemic therapies are primarily indicated for advanced-stage tumors that have metastasized the lymph nodes (N2-N4). In contrast, N0 / N1 tumors are typically managed with surgical resection, as surgery remains the standard curative approach for these early-stage cases ^23^.

However, the potential of anti-angiogenic therapy has yet to be fully demonstrated, due to numerous limitations and resistance mechanisms ^19^. Moreover, the prevailing hypothesis that tumor growth depends on angiogenesis has been challenged by the description of a non-angiogenic growth ^21^. This non-angiogenic growth, termed vessel co-option, involves tumor cell proliferation without compromising parenchymal integrity, thereby preserving the original structure and enabling hijacking of existing blood vessels rather than creating new ones ^20^. This process has been identified as an additional mechanism of tumor vascularization and is considered a resistance mechanism to anti-angiogenic therapy ^10^. Consequently, the hallmark ‘Inducing angiogenesis’ has been replaced by ‘Inducing or accessing vasculature’ in the 2022 revision of the ‘Hallmarks of Cancer’ ^24^.

Identifying tumor vascularization mechanisms is crucial for determining which patients are most likely to respond to anti-angiogenic therapies. This approach could increase response rates while avoiding side effects in non-responsive patients. Moreover, determining the tumor vascularization pattern also has important prognostic implications. While angiogenic tumors have traditionally been considered more aggressive and invasive, some authors have challenged this idea, proposing that within the same histological subtype, co-option tumors are more aggressive than angiogenic tumors ^5,21^, presenting a less favorable microenvironment.

In the present study, samples obtained from untreated N0 / N1 stage tumors were analyzed ^23^. This approach enabled assessment of tumor development unaffected by therapeutic interventions, providing valuable insights into intrinsic histopathological and vascular characteristics of early-stage NSCLC.

The identification of the vascularization mechanism was based on the spatial analysis of tumor microvasculature. Typical markers used to assess the density of the microvasculature (MVD), such as the endothelial markers CD34 or CD31, are generally employed for this purpose ^6,25,26^. Most authors describe vessel co-option as being characterized by the preservation of the distinctive pattern of normal lung tissue, exhibiting a honeycomb-like structure, in contrast to the destructive pattern associated to angiogenesis ^4,27^. Using CD31 labeling, most samples were successfully identified (76.6%), while 25 samples remained unidentified or mixed. Furthermore, some authors have proposed a classification of tumors according to whether they present a destructive pattern with formation of new parenchyma, which is associated with the angiogenic pattern, or a non-destructive pattern ^12,28,29^. Based on this observation, a second staining using WVG technique was performed to enhance identification success rates, a second staining was performed using WVG technique. WVG stain identifies collagen in red color and elastin fibers in purple; therefore, the criterion used to identify parenchymal destruction was the presence of intact or fragmented elastin. With this combined approach, we confirmed that many samples, mainly papillary and micropapillary, presented a mixed pattern, characterized by the coexistence of angiogenic and co-option patterns. Considering these truly mixed samples, we completed the classification of the 25 ambiguous samples. However, a number of samples were identified that presented conflicting classifications depending on the staining method used. That is, there were samples that were initially considered to be a type of vascular pattern, but the second staining cast doubt on this classification. Despite the limited number of samples, we believe that it is relevant to study them in depth since they may be identifying a transition stage between one pattern and another. Consequently, our dual approach has enhanced the efficiency of vascular pattern identification by 20% when compared to CD31 staining alone.

It is evident that conducting this double determination (CD31 and WVG) in biopsies from patients with metastatic NSCLC (N2-N4) would facilitate a more precise adjustment of the therapeutic approach. However, it is difficult to perform specific stains to identify vascularization, as they are not part of routine hospital practice. Additionally, biopsy samples are highly valuable, as they are typically obtained from patients in advanced stages and are often small, with priority given to histological and molecular diagnosis. In this context, our data offer valuable information as they reveal correlations between specific histopathologic and vascular patterns. Specifically, the solid basal and diffuse patterns correlate perfectly with an angiogenic pattern, while the solid alveolar pattern corresponds to vascular co-option. The lepidic pattern, which is associated with a distinct vascular pattern, in most cases corresponds to vessel co-option. As for the papillary and micropapillary patterns, they mostly showed a mixed vascular pattern, with the co-option pattern presented in the fibrovascular septa and the angiogenic pattern in the tufts. Interestingly, papillary tumors that were not mixed, were classified as co-option, while micropapillary tumors were angiogenic. The acinar pattern is the most complex, given that although most are angiogenic with a destructive pattern, a relevant percentage of co-option tumors is observed, and this is where the unclassified samples are included. These findings align with relationships proposed by previous studies ^4,6,28^. However, it should be noted that these studies predate the new WHO classification of NSCLC growth patterns and, as such, are incomplete at this time. To address this limitation, we have undertaken a comprehensive study, involving the correlation between the newly described patterns and the vascularization patterns, utilizing a larger sample.

Consequently, our results add further value to histologic analysis of biopsies, which remains relevant despite the growth of molecular diagnostics. Just as numerous treatable molecular alterations are associated with specific histological subtypes ^13^, this study demonstrates that certain tumor vascularization mechanisms are associated to specific histopathologic subtypes. This finding may have important therapeutic implications, especially when the tumor stage is at least N2, as it suggests that anti-angiogenic therapies may be less effective for tumors identified as solid alveolar or lepidic and may be more effective in tumors classified as basal or diffuse solid. For papillary or micropapillary tumors, some response to therapy could be expected, but a portion of the tumor may remain resistant to therapy and a change in the growth pattern could be induced. Finally, our findings may have prognostic implications, since in subtypes where both angiogenic and co-option vascularization patterns are found, the latter could have a worse prognosis than the former ^5,21^.

### CRediT author statement

**Inés Solano-SC:** Conceptualization, Methodology, Investigation, Visualization, Writing - Original Draft. **Marta Rodríguez-González:** Validation, Writing - Review & Editing. **María González-Núñez:** Supervision, Writing - Review & Editing. **Alicia Rodríguez-Barbero:** Project administration, Writing - Review & Editing. **José Manuel Muñoz-Félix:** Conceptualization, Methodology, Supervision, Project administration, Funding acquisition, Writing - Original Draft. **Miguel Pericacho:** Conceptualization, Methodology, Visualization, Supervision, Project administration, Funding acquisition, Writing - Review & Editing.

## Data Availability

All data produced in the present study are available upon reasonable request to the authors

## Acknowledgements

This research was funded by the Ministerio de Economía y Competitividad of Spain (PID2022-138765OB-I00 to MP), the Instituto de Salud Carlos III (PI19/01630 and co-funded by FEDER to MP and PI21/01034 to JMM-F), Fundación Científica de la Asociación Española Contra el Cáncer (LABAE234462MUÑO to JMM-F), Fundación Mutua Madrileña (Ayudas a la Investigación en Salud 2024 to JMM-F) and Junta de Castilla y León (BIO/SA83/13 to MP and SA095P24 to JMM-F).

Furthermore, IS-SC were supported by a contract from the Junta de Castilla y León (co-funded by the European Social Fund).

## Declaration of generative AI and AI-assisted technologies in the writing process

During the preparation of this work the authors used DeepL and Perplexity in order to improve language and readability. After using these tools, the authors reviewed and edited the content as needed and take full responsibility for the content of the publication.

---

**Supplementary Figure 1:**
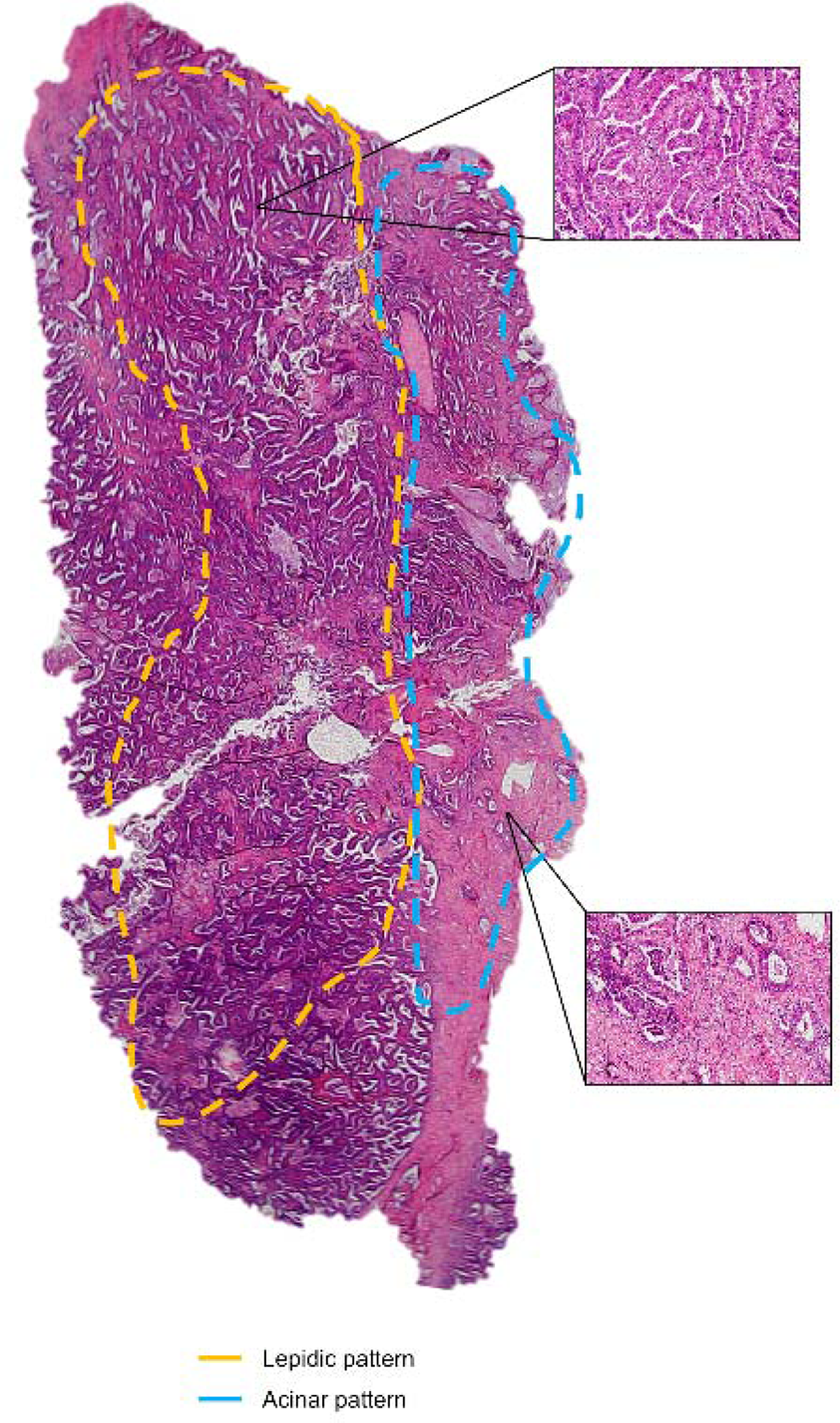
Whole sample analyzed by delineating different patterns as described by Yoshizawa^22^ using hematoxylin/eosin staining. A sample exhibited a 50% lepidic growth pattern and a 50% of acinar pattern (sample number #5 in Supplementary Table 1).

**Supplementary Figure 2:**
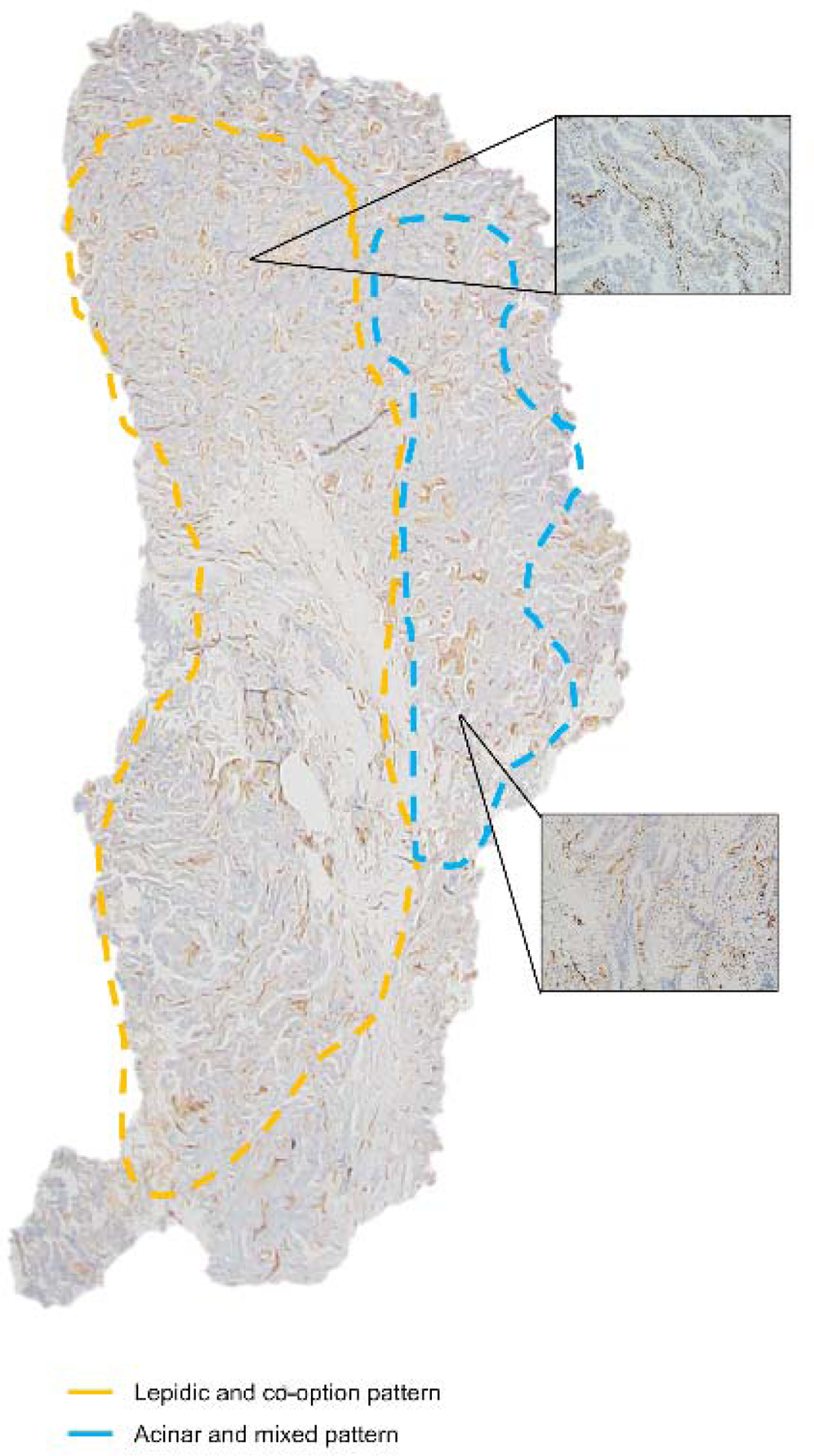
Whole sample analyzed by delimiting different vascular patterns like angiogenic or non-angiogenic by CD31 staining. An example of a sample with some mixed vascular pattern in blue and with non-angiogenic pattern in orange (sample number #5 in Supplementary Table 1).

**Supplementary Figure 3:**
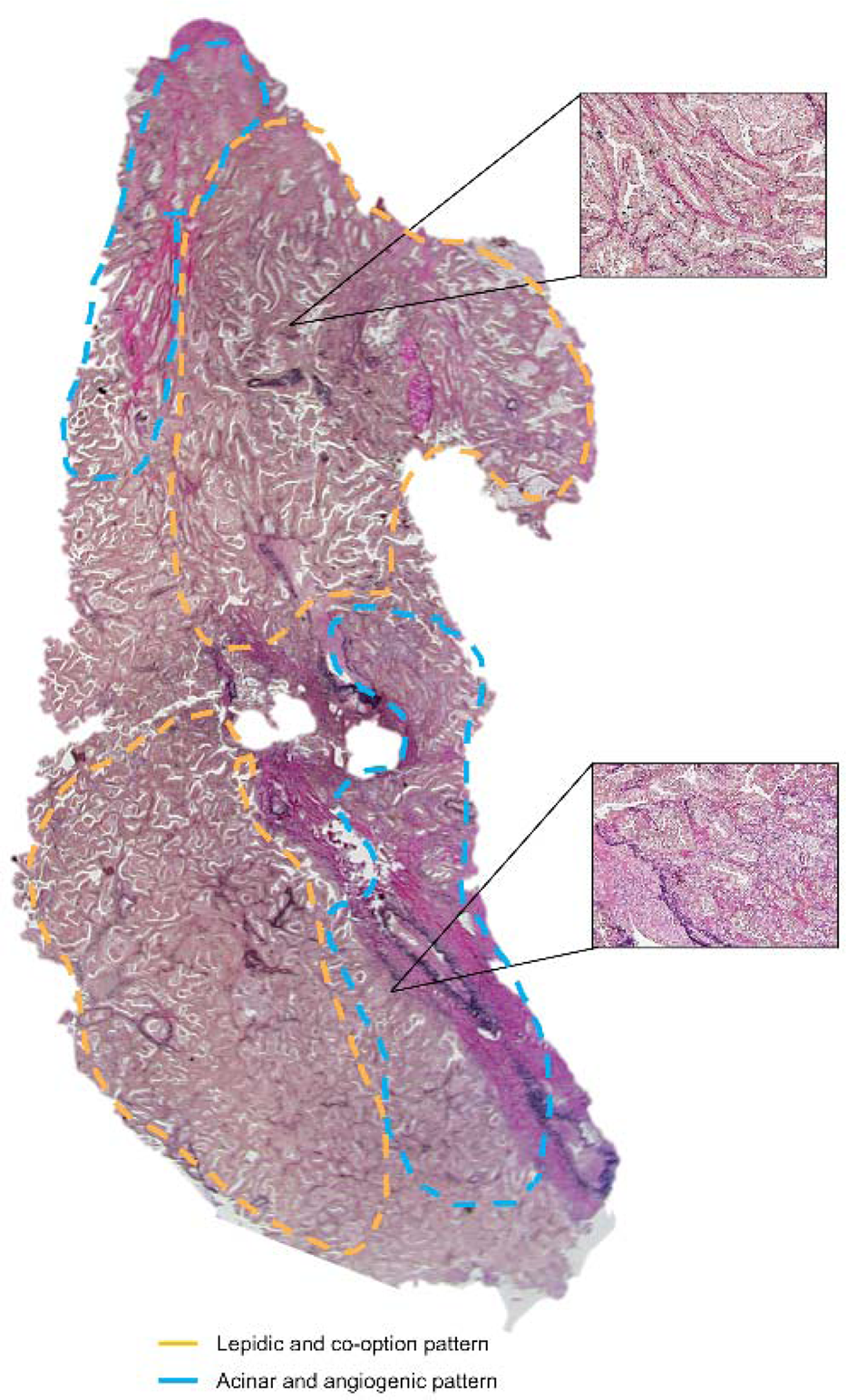
Whole sample analyzed by delimiting different vascular patterns like angiogenic or non-angiogenic by Weigert-Van Gieson staining. An example of a sample with some angiogenic patterns in blue and with non-angiogenic pattern in orange (sample number #5 in Supplementary Table 1).

**Supplementary Table 1:** Histopathological and vascular classification of 70 samples of human lung adenocarcinoma. A: Angiogenic; C: Co-option; M: Mixed

| ID | HEMATOXYLIN/EOSIN | CD31 | WVG | General |
| --- | --- | --- | --- | --- |
| #1 | Diffuse solid | A | A | A |
| #2 | - Acinar (45%)<br>- Micropapillary (45%)<br>- Lepidic (10%) | - Acinar: A<br>- Micropapillary: A<br>- Lepidic: C | - Acinar: A-M<br>- Micropapillary: A<br>- Lepidic: C | - Acinar: A<br>- Micropapillary: A<br>- Lepidic: C |
| #3 | Alveolar solid | C | C | C |
| #4 | Acinar | A-M | M | M |
| #5 | - Lepidic (50%)<br>- Acinar (50%) | - Lepidic: C<br>- Acinar: M-C | - Lepidic: C<br>- Acinar: A-M | - Lepidic: C<br>- Acinar: M |
| #6 | - Acinar (40%)<br>- Lepidic (60%) | - Acinar: M<br>- Lepidic: C | - Acinar: C<br>- Lepidic: C | - Acinar: C<br>- Lepidic: C |
| #7 | - Acinar (60%)<br>- Lepidic (40%) | - Acinar: C<br>- Lepidic: C | - Acinar: C<br>- Lepidic: C | - Acinar: C<br>- Lepidic: C |
| #8 | Acinar | Acinar: A-M | Acinar: A-M | Acinar: A |
| #9 | Basal solid | A-M | A | A |
| #10 | - Diffuse solid (60%)<br>- Acinar (30%)<br>- Alveolar solid (10%) | - Diffuse solid: A<br>- Acinar: C<br>- Alveolar solid: C | - Diffuse solid: A<br>- Acinar: M<br>- Alveolar solid: C | - Diffuse solid: A<br>- Acinar: C<br>- Alveolar solid: C |
| #11 | Acinar | A | A | A |
| #12 | - Papillary (50%)<br>- Acinar (40%)<br>- Lepidic (10%) | - Papillary: M<br>- Acinar: A<br>- Lepidic: M | - Papillary: C-M<br>- Acinar: A-M<br>- Lepidic: C | - Papillary: M<br>- Acinar: A<br>- Lepidic: C |
| #13 | Acinar | A | A | A |
| #14 | Lepidic | C | C | C |
| #15 | Acinar | C | C | C |
| #16 | Solid:<br>- Alveolar (70%)<br>- Diffuse (30%) | - Alveolar: C<br>- Diffuse: A-M | - Alveolar: M<br>- Diffuse: A-M | - Alveolar: C<br>- Diffuse: A |
| #17 | - Acinar (60%)<br>- Lepidic (40%) | - Acinar: A<br>- Lepidic: M | - Acinar: C<br>- Lepidic: C | - Acinar: UC<br>- Lepidic: C |
| #18 | Acinar | A | M | A |
| #19 | - Acinar (85%)<br>- Basal solid (15%) | - Acinar: A<br>- Basal solid: A | - Acinar: A<br>- Basal solid: A | - Acinar: A<br>- Basal solid: A |
| #20 | Acinar | A | A | A |
| #21 | - Papillary (85%)<br>- Lepidic (15%) | - Papillary: C-M<br>- Lepidic: C | - Papillary: C-M<br>- Lepidic: C | - Papillary: C<br>- Lepidic: C |
| #22 | Acinar | C | A | UC |
| #23 | Acinar (cribiform) | C-M | C | C |
| #24 | - Lepidic (50%)<br>- Acinar (50%) | - Lepidic: C<br>- Acinar: A | - Lepidic: C<br>- Acinar: C | - Lepidic: C<br>- Acinar: UC |
| #25 | Acinar | A | A | A |
| #26 | - Lepidic (15%)<br>- Acinar (85%) | - Lepidic: A<br>- Acinar: A-M | - Lepidic: A<br>- Acinar: A | - Lepidic: A<br>- Acinar: A |
| #27 | Micropapillary | M | M | M |
| #28 | - Basal solid (60%)<br>- Acinar (40%) | - Basal solid: A<br>- Acinar: C | - Basal solid: A<br>- Acinar: A | - Basal solid: A<br>- Acinar: UC |
| #29 | Diffuse solid | A | A | A |
| #30 | Acinar | A | C | UC |
| #31 | Acinar | M | A | A |
| #32 | - Lepidic (50%)<br>- Acinar (50%) | - Lepidic: C<br>- Acinar: M | - Lepidic: C<br>- Acinar: C | - Lepidic: C<br>- Acinar: C |
| #33 | Acinar | A | A | A |
| #34 | Acinar | C | C | C |
| #35 | Acinar | A | A | A |
| #36 | Lepidic | A | A | A |
| #37 | Acinar | A-M | A | A |
| #38 | - Papillary (30%)<br>- Acinar (60%)<br>- Alveolar solid (10%) | - Papillary: C<br>- Acinar: A<br>- Alveolar solid: C | - Papillary: C<br>- Acinar: M<br>- Solid: C | - Papillary: C<br>- Acinar: A<br>- Alveolar solid: C |
| #39 | - Papillary (90%)<br>- Acinar (10%) | - Papillary: M<br>- Acinar: M | - Papillary: M-C<br>- Acinar: C | - Papillary: M<br>- Acinar: C |
| #40 | - Diffuse solid (50%)<br>- Acinar (35%)<br>- Alveolar solid (15%) | - Diffuse solid: A<br>- Acinar: A<br>- Alveolar solid: C | - Diffuse solid: A<br>- Acinar: A<br>- Alveolar solid: C | - Diffuse solid: A<br>- Acinar: A<br>- Alveolar solid: C |
| #41 | Acinar | A | A | A |
| #42 | Acinar | A | C | UC |
| #43 | Solid:<br>- Alveolar (60%)<br>- Diffuse (40%) | - Alveolar: C<br>- Diffuse: A | - Alveolar: M<br>- Diffuse: A | - Alveolar: C<br>- Diffuse: A |
| #44 | - Acinar (90%)<br>- Micropapillary (10%) | - Acinar: A<br>- MP: M | - Acinar: A<br>- MP: M | - Acinar: A<br>- MP: M |
| #45 | Acinar | A | A | A |
| #46 | - Diffuse solid (50%)<br>- Acinar (50%) | - Solid: A<br>- Acinar: A | - Solid: A<br>- Acinar: A | - Solid: A<br>- Acinar: A |
| #47 | - Alveolar solid (50%)<br>- Acinar (50%) | - Alveolar solid: A<br>- Acinar: A | - Alveolar solid: A<br>- Acinar: A | - Alveolar solid: A<br>- Acinar: A |
| #48 | Alveolar solid | C | M | C |
| #49 | Solid:<br>- Alveolar (40%)<br>- Diffuse (60%) | - Alveolar: C<br>- Diffuse: A-M | - Alveolar: C<br>- Diffuse: A | - Alveolar: C<br>- Diffuse: A |
| #50 | Diffuse solid | A | A-M | A |
| #51 | - Diffuse solid (20%)<br>- Acinar (70%)<br>- Lepidic (10%) | - Solid: A<br>- Acinar: C<br>- Lepidic: M | - Solid: A<br>- Acinar: C<br>- Lepidic: C | - Solid: A<br>- Acinar: C<br>- Lepidic: C |
| #52 | Solid:<br>- Basal (50%)<br>- Diffuse (50%) | - Basal: A<br>- Diffuse: A | - Basal: A<br>- Diffuse: A | - Basal: A<br>- Diffuse: A |
| #53 | Acinar | A-M | C-M | M |
| #54 | Acinar | A | A | A |
| #55 | Basal solid | A | A | A |
| #56 | Acinar | A | A-M | A |
| #57 | Alveolar solid | C | C | C |
| #58 | Acinar | A | A | A |
| #59 | Acinar | A | C | UC |
| #60 | Acinar | A | A | A |
| #61 | Basal solid | A | A | A |
| #62 | Basal solid | A | A | A |
| **#63** | Diffuse solid | A | A-M | A |
| **#64** | Solid<br>- Alveolar (50%)<br>- Diffuse (50%) | - Alveolar: C<br>- Diffuse: A | - Alveolar: M<br>- Diffuse: A | - Alveolar: C<br>- Diffuse: A |
| **#65** | Diffuse solid | A | A | A |
| **#66** | - Lepidic (60%)<br>- Acinar (40%) | - Lepidic: C<br>- Acinar: C | - Lepidic: C<br>- Acinar: C | - Lepidic: C<br>- Acinar: C |
| **#67** | Lepidic | C | C | C |
| **#68** | - Acinar (40%)<br>- Papillary (40%)<br>- Lepidic (20%) | - Acinar: A<br>- Papillary: A-M<br>- Lepidic: M | - Acinar: A<br>- Papillary: M<br>- Lepidic: C | - Acinar: A<br>- Papillary: M<br>- Lepidic: C |
| **#69** | - Papillary (50%)<br>- Micropapillary (50%) | M | M | M |
| **#70** | Acinar | A | A | A |

## BIBLIOGRAPHY

1. Sung H, Ferlay J, Siegel RL, et al. Global Cancer Statistics 2020: GLOBOCAN Estimates of Incidence and Mortality Worldwide for 36 Cancers in 185 Countries. CA: A Cancer Journal for Clinicians. 2021;71(3):209–249. doi:10.3322/caac.21660

2. Herbst RS, Morgensztern D, Boshoff C. The biology and management of non-small cell lung cancer. Nature. 2018;553(7689):446–454. doi:10.1038/nature25183

3. WHO Classification of Tumours Editorial Board, ed. Thoracic Tumours. 5th ed. International Agency for Research on Cancer; 2021.

4. Kuczynski EA, Vermeulen PB, Pezzella F, Kerbel RS, Reynolds AR. Vessel co-option in cancer. Nat Rev Clin Oncol. 2019;16(8):469–493. doi:10.1038/s41571-019-0181-9

5. Paulsen EE, Andersen S, Rakaee M, et al. Impact of microvessel patterns and immune status in NSCLC: a non-angiogenic vasculature is an independent negative prognostic factor in lung adenocarcinoma. Front Oncol. 2023;13:1157461. doi:10.3389/fonc.2023.1157461

6. Pezzella F, Pastorino U, Tagliabue E, et al. Non-Small-Cell Lung Carcinoma Tumor Growth without Morphological Evidence of Neo-Angiogenesis. 1997;151(5).

7. Kuhn E, Morbini P, Cancellieri A, Damiani S, Cavazza A, Comin CE. Adenocarcinoma classification: patterns and prognosis. Pathologica. 2018;110(1):5–11.

8. Russell PA, Wainer Z, Wright GM, Daniels M, Conron M, Williams RA. Does Lung Adenocarcinoma Subtype Predict Patient Survival?: A Clinicopathologic Study Based on the New International Association for the Study of Lung Cancer/American Thoracic Society/European Respiratory Society International Multidisciplinary Lung Adenocarcinoma Classification. Journal of Thoracic Oncology. 2011;6(9):1496–1504. doi:10.1097/JTO.0b013e318221f701

9. Yoshizawa A, Motoi N, Riely GJ, et al. Impact of proposed IASLC/ATS/ERS classification of lung adenocarcinoma: prognostic subgroups and implications for further revision of staging based on analysis of 514 stage I cases. Modern Pathology. 2011;24(5):653–664. doi:10.1038/modpathol.2010.232

10. Kuczynski EA, Reynolds AR. Vessel co-option and resistance to anti-angiogenic therapy. Angiogenesis. 2020;23(1):55–74. doi:10.1007/s10456-019-09698-6

11. De Oliveira Duarte Achcar R, Nikiforova MN, Yousem SA. Micropapillary Lung Adenocarcinoma. American Journal of Clinical Pathology. 2009;131(5):694–700. doi:10.1309/AJCPBS85VJEOBPDO

12. Donnem T, Reynolds AR, Kuczynski EA, et al. Non-angiogenic tumours and their influence on cancer biology. Nat Rev Cancer. 2018;18(5):323–336. doi:10.1038/nrc.2018.14

13. Conde E, Angulo B, Izquierdo E, et al. Lung adenocarcinoma in the era of targeted therapies: histological classification, sample prioritization, and predictive biomarkers. Clin Transl Oncol. 2013;15(7):503–508. doi:10.1007/s12094-012-0983-z

14. Von Der Thüsen JH, Tham YS, Pattenden H, et al. Prognostic Significance of Predominant Histologic Pattern and Nuclear Grade in Resected Adenocarcinoma of the Lung: Potential Parameters for a Grading System. Journal of Thoracic Oncology. 2013;8(1):37–44. doi:10.1097/JTO.0b013e318276274e

15. Ngaha TYS, Zhilenkova AV, Essogmo FE, et al. Angiogenesis in Lung Cancer: Understanding the Roles of Growth Factors. Cancers. 2023;15(18):4648. doi:10.3390/cancers15184648

16. Weis SM, Cheresh DA. Tumor angiogenesis: Molecular pathways and therapeutic targets. Nature Medicine. 2011;17(11):1359–1370. doi:10.1038/nm.2537

17. Vasudev NS, Reynolds AR. Anti-angiogenic therapy for cancer: current progress, unresolved questions and future directions. Angiogenesis. 2014;17(3):471–494. doi:10.1007/s10456-014-9420-y

18. Hall RD. Angiogenesis inhibition as a therapeutic strategy in non-small cell lung cancer (NSCLC). Translational lung cancer research. 2015;4(5).

19. Haibe Y, Kreidieh M, El Hajj H, et al. Resistance Mechanisms to Anti-angiogenic Therapies in Cancer. Front Oncol. 2020;10:221. doi:10.3389/fonc.2020.00221

20. Seano G, Jain RK. Vessel co-option in glioblastoma: emerging insights and opportunities. Angiogenesis. 2020;23(1):9–16. doi:10.1007/s10456-019-09691-z

21. Sardari Nia P, Colpaert C, Blyweert B, et al. Prognostic value of nonangiogenic and angiogenic growth patterns in non-small-cell lung cancer. Br J Cancer. 2004;91(7):1293–1300. doi:10.1038/sj.bjc.6602134

22. Yoshizawa A, Sumiyoshi S, Sonobe M, et al. Validation of the IASLC/ATS/ERS Lung Adenocarcinoma Classification for Prognosis and Association with EGFR and KRAS Gene Mutations: Analysis of 440 Japanese Patients. Journal of Thoracic Oncology. 2013;8(1):52–61. doi:10.1097/JTO.0b013e3182769aa8

23. Postmus PE, Kerr KM, Oudkerk M, et al. Early and locally advanced non-small-cell lung cancer (NSCLC): ESMO Clinical Practice Guidelines for diagnosis, treatment and follow-up. Annals of Oncology. 2017;28:iv1–iv21. doi:10.1093/annonc/mdx222

24. Hanahan D. Hallmarks of Cancer: New Dimensions. Cancer Discovery. 2022;12(1):31–46. doi:10.1158/2159-8290.CD-21-1059

25. Damianovich M, Hout Siloni G, Barshack I, et al. Structural Basis for Hyperpermeability of Tumor Vessels in Advanced Lung Adenocarcinoma Complicated by Pleural Effusion. Clinical Lung Cancer. 2013;14(6):688–698. doi:10.1016/j.cllc.2013.06.007

26. Vermeulen P, Pezzella F. Nonangiogenic tumor growth. In: Tumor Vascularization. Elsevier; 2020:15–32. doi:10.1016/B978-0-12-819494-2.00002-X

27. Bridgeman VL, Vermeulen PB, Foo S, et al. Vessel co-option is common in human lung metastases and mediates resistance to anti-angiogenic therapy in preclinical lung metastasis models. J Pathol. 2017;241(3):362–374. doi:10.1002/path.4845

28. Sardari Nia P, Colpaert C, Vermeulen P, et al. Different Growth Patterns of Non-Small Cell Lung Cancer Represent Distinct Biologic Subtypes. The Annals of Thoracic Surgery. 2008;85(2):395–405. doi:10.1016/j.athoracsur.2007.08.054

29. Suzuki S, Aokage K, Hishida T, et al. Interstitial growth as an aggressive growth pattern in primary lung cancer. J Cancer Res Clin Oncol. 2016;142(7):1591–1598. doi:10.1007/s00432-016-2168-6

